# REVISITING TUBERCULOSIS (TB) DIAGNOSIS: THE PROSPECT OF URINE LIPOARABINOMANNAN (LAM) ASSAY AS A DIAGNOSTIC TOOL IN PEOPLE LIVING WITH HIV

**DOI:** 10.1101/2024.10.09.24315199

**Authors:** Ayodele Isaac Adedokun, Gaius Bala, Nafisa M.K. Elehamer, Raphael Yohanna, Boluwatife Deborah Oyelabi, Samuel Eniola Gana

**Affiliations:** Department of Chemical Pathology, School of Medical Laboratory Science, Usmanu Danfodiyo University Sokoto, Nigeria; Department of Histopathology, School of Medical Laboratory Science, Usmanu Danfodiyo University Sokoto, Nigeria; Department of Public Health and Epidemiology, Faculty of Medicine, University of Debrecen, Debrecen, Hungary; Faculty of Public and Environmental Health, University of Khartoum, Khartoum, Sudan; Department of Nursing Science, Faculty of Basic Medical Science, Osun State University, Nigeria; Department of Immunology, School of Medical Laboratory Science, Usmanu Danfodiyo University Sokoto, Nigeria

**Keywords:** Tuberculosis, Lipoarabinomannan, HIV, diagnosis, urine, global health

## Abstract

**Background:** Tuberculosis (TB), since its discovery has become a global health burden, continuously spreading across the nations of the earth with increased mortality. Its effect on human immunodeficiency virus (HIV) progression and immune cell deterioration cannot be overlooked. The widespread use of traditional and molecular TB diagnostic methods has been hindered by several factors. Nonetheless, the development of a rapid, sensitive, timely, non-invasive diagnostic protocol could revolutionize the diagnostic era and halt the spread of this infectious agent. It could also help in the early diagnosis of the infection in poorly developed areas. The detection efficacy of urine lipoarabinomannan assay as a prospect in tuberculosis diagnosis in HIV-positive patients was assessed in this study. Urine samples were collected into a wide-mouthed universal container from HIV-positive patients with signs and symptom of tuberculosis and HIV advanced disease. Alere Determine urine lateral flow - LAM test was used for the detection of LAM antigen in urine samples of HIV-positive patients. The CD4 count of the HIV-positive patients was also documented.

**Results:** A total of 85 HIV-positive subjects were included in the analysis based on the inclusion criteria. Urinary LAM was reactive in 29 (34.1%) of 85 HIV-positive patients and non-reactive in 56 (69.1%). The majority of the participants had a CD4^+^ count < 200 cells/µL 60 (70.6%) while others had CD4^+^ count ≥ to 200 cells/µL 25 (29.4%). There was no statistically significant difference between CD4 count and LAM positivity. Most of the affected age groups in the study were 31-45 years, and females had preponderance in comparison to male subjects.

**Conclusions:** The use of urinary LAM in HIV-positive patients is a helpful tool for TB screening. It also has a promising ability for early TB diagnosis in the study group and other TB-associated disease conditions in resource-deprived settings. For the general populace, further research is required to certify its sensitivity and specificity.

## 1. Background

In the history of disease intervention, tuberculosis is a global health pandemic that has caused the lives of millions over the past centuries [1]. Caused by *Mycobacterium tuberculosis* (MTB), this airborne transmitted pathogen is responsible for 4300 deaths/day, making it the second most destructive in the world; led by the COVID-19 virus [2]. Among all the TB cases documented globally through World Health Organization (WHO) reports, 6.3% are observed in people living with human immunodeficiency virus (PLWH) compared to those without the disease [3,4]. Data from the WHO in 2022 estimated the number of incident TB cases, for nations with a minimum of 100,000 incident cases accounting for about two-thirds of global cases, specifically eight countries in ascending order. Nigeria ranked 6th globally and was the most burdened country in Africa [4]. The prevalence of TB, including drug-resistant TB, in different locations in Nigeria has been reported to be 5.8%, 9.9%, 13.8% and 16.2% [5–7]. Disruptions in TB diagnosis and treatment due to the COVID-19 pandemic closely correlate with these increases in TB incidence [4].

For TB prevention, diagnosis, and treatment, a rapid and accurate testing protocol is essential to prevent further spread of bacteria [8]. Various diagnostic techniques, including biomolecular, immunological, radiological, microscopic, bacterial culture, and clinical methods, are currently being employed [8]. Molecular tests such as geneXpert, ELISA, and line probe assays are currently available to reform the TB diagnostic landscape and augment the current conventional methods [4,9]. However, their widespread use is often hindered by operator efficiency, costs, unavailability of laboratory resources, power supply, and low sensitivity in populations such as children with HIV [10]. Certain factors, such as poverty, inefficient healthcare service delivery, lack of patient compliance, and follow-up, have also contributed to the emergence and re-emergence of drug-resistant (DR) strains [11].

Frequently, conventional disease diagnosis includes invasive methods that are usually painful, anxiety-bound, and traumatic. Nevertheless, progress in the identification of non-invasive biomarkers has created exciting opportunities through the use of easily accessible bodily fluids that can provide information on health and disease [12]. Among all known bodily fluids, urine poses as a great candidate that can revamp the diagnostic era. It offers various advantages ranging from convenient non-invasive specimen collection, cost-effectiveness, variety of biomarker molecules, and reasonable sample volume for multiple diagnostic testing [12]. The potential of urine is becoming more evident for the diagnosis and follow-up of various diseases, particularly infectious diseases ravaging low- and middle-income countries [13,14].

Recently, organisations such as the National Institute of Allergy and Infectious Diseases (NIAID) are working to strengthen advances in the feasibility of novel diagnostics of TB in Endemic Countries (FEND TB), through programs that appreciate early stage diagnostics in endemic areas [15]. Active research into developing accurate, sensitive, and timely methods of TB prevention, including screening, diagnosis, treatment, and development of effective vaccines, is ongoing. Research has shown that most individuals with presumptive cases are unable to produce sputum of high- quality and volume. A non-sputum-based protocol with increased turnaround time could serve as an alternative [16].

The Alere Determine urine lateral flow (LF) - lipoarabinomannan (LAM) assay is an immunocapture point-of-care test that detects mycobacterial LAM antigens in urine [17]. It is the WHO recommended urine test for TB diagnosis [18,19]. The use of the LAM bacterial antigen, which is an integral constituent of mycobacterial cell wall [20], is associated with its heat stability. This heat-stable antigen is released from metabolically active or degrading Mycobacteria [16] and is filtered by the kidneys into the urine. LAM is recognised for its ability to induce anti-inflammatory effects on the immune system by suppressing both innate and adaptive immunity while simultaneously enhancing the release of IL-10 [20]. This effect could impact other immune cells, such as CD4^+^ T- cells, macrophages and B- cells.

In this study, we assessed the diagnostic efficacy of the urine-based AlereLAM test as a rule-in screening test for early TB detection in patients with HIV immunosuppression and the impact of LF-LAM implementation on TB mortality in resource-deprived settings. We analysed the recency status of HIV-positive patients with reactive and non-reactive LAM results.

## 2 Methods

### 2.1 Study design and population

This study included PLWH assessing the medical laboratory unit of a general hospital facility under the Ministry of Health in Akwa Ibom State, Nigeria. This study complied with the requirements for laboratory testing standards. As tuberculosis is a major health concern in our country, this study proves societal importance and global health coverage. Furthermore, all diagnostic procedures were performed by experienced healthcare professionals. Hospital numbers, CD4 count, HIV confirmatory status, and demographic data were stored and disposed of after the study timeline. The study timeline was 3 months from onset to completion of the study.

GeneXpert assay, sputum culture, and microscopy were not performed in tandem during this study because of the unavailability of resources to carry out testing in the healthcare facility. In this study, we also determined the recency status. Reports show that the population area of the study has one of the highest incidences of HIV (3.5%) in Nigeria [21], and considering these settings will help make more important statistical reconciliations and infer population-based drift in HIV information. The methodology adhered to the ethical criteria outlined in the 1975 Declaration of Helsinki and received approval from the Medical Ethics Committee of the Healthcare Centre. We obtained informed consent from patients prior to screening (MOH/GHUA/2024/006).

### 2.2 Inclusion and Exclusion Criteria

This study included any HIV-positive patient who satisfied one of the following criteria for participation: (1) TB symptoms including chills, cough, fever, night sweating, and greater than 18 years of age (2) advanced HIV disease. Patients who had already been placed on anti-TB therapy, < 18 years old, or HIV-negative patients were excluded from this study. The flowchart for the diagnostic algorithm is shown in (Fig. 1).

**Fig. 1.**
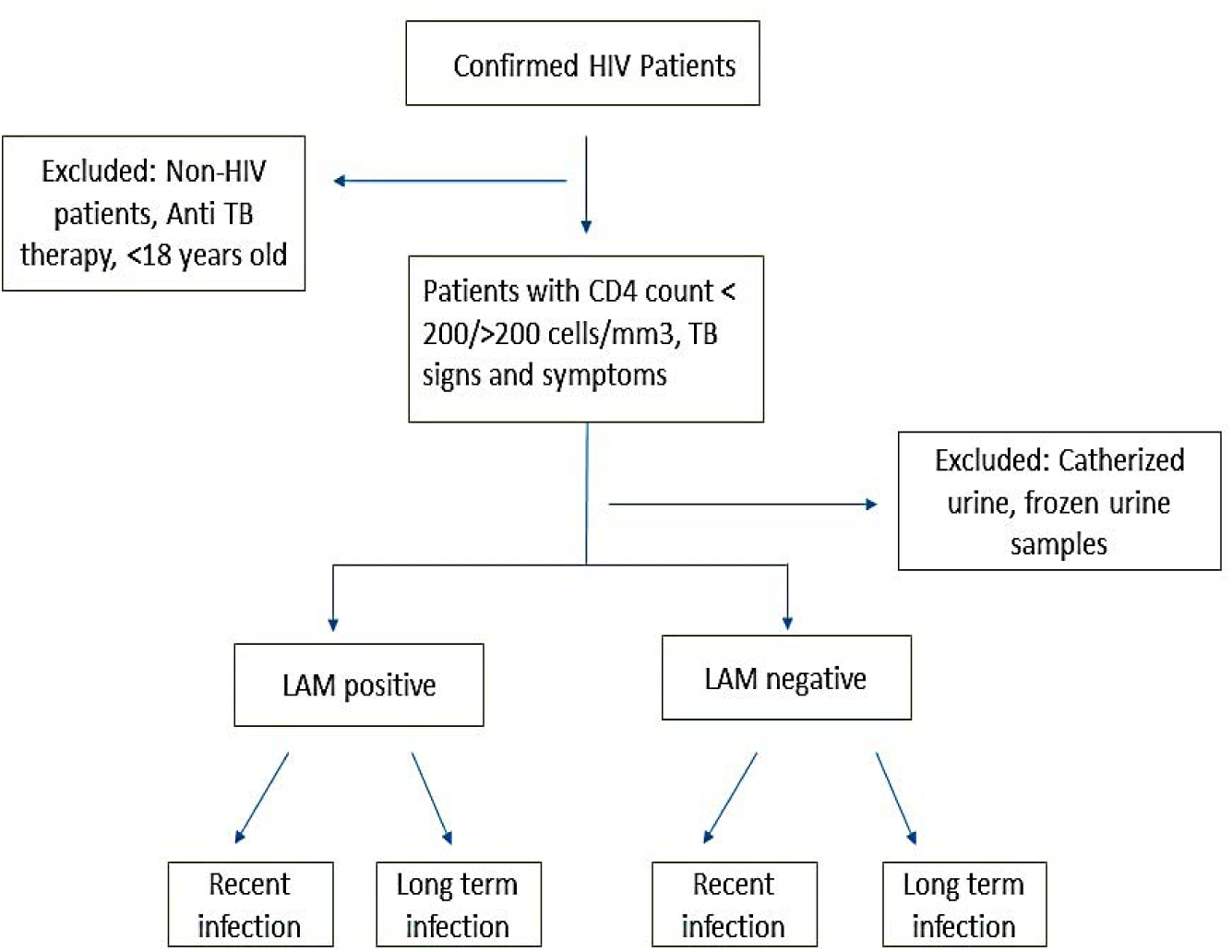
Flowchart diagram for the diagnostic testing.

### 2.3 Sample collection

#### 2.3.1 Urine collection

Random urine samples were collected in a labelled sterile universal wide-mouthed urine container with a minimum volume of 5 mL. After the sample collection, the samples were labelled appropriately and stored at room temperature, and testing was completed within 2 hours of collection. The samples collected were neither frozen nor collected through a catheter.

### 2.4 Laboratory analysis

#### 2.4.1 LAM testing

Alere Determine TB LAM Ag lateral flow assay test strips (Abbott Laboratories, Lake Bluff, USA) was used to determine urinary LAM. 60 µL of urine sample was added to the sample pot area using a micropipette. After 25 minutes, the result was read and interpreted according to the manufacturer’s instructions, using the manufacturer’s reference card to determine if a test was valid, and then to determine if the test result was reactive or not. A test was recorded as reactive if a visible line was noted in the “test” and “control” areas of the test strip that matched the manufacturer’s reference card. A test without a line on the “patient area” but on the “control” area was deemed non-reactive.

### 2.5 Statistical analysis

Data were entered into Microsoft Excel 2019. The data were analysed using SPSS 26.0 statistical software (SPSS Inc., Chicago, IL). The frequencies, mean, and median values were calculated. The chi-squared test was used for categorical variables. p-Values of less than 0.05 were considered statistically significant.

## 3 Results

### 3.1 Demographic characteristics of the study population

130 HIV-positive patients were assessed in the laboratory unit for clinical investigation during the study period. A total of 85 HIV-positive patients were enrolled in the study based on the inclusion criteria. Females have a preponderance in comparison to males. 65 of the participants were females (76.5%), and 20 of the study participants were males (23.5%). The maximum number of participants belonged to the age group 31-45 with a percent of 46 (54.1%), followed by the age group 15-30 (35.3%) with a mean age of 35 (SD± 10.19).

The CD4^+^ cell count was also determined in all the HIV-positive patients with presumptive TB symptoms in the study. In total, 60 of 85 (70.6%) HIV-positive patients had a CD4^+^ count < 200 cells/µL, while 25 of 85 (29.4%) HIV-positive patients had a CD4^+^ count ≥ 200 cells/µL. The median CD4^+^ count was 100 cells/µL. The demographic and clinical details of the patients as described show that the highest number of patients with CD4^+^ count < 200 cells/µL (56.7%) and CD4^+^ count ≥ to 200 cells/µL (48.0%) belonged to the age group of 31-45 years, followed by the age group 15-30. This also applies to sex in which females had a lower CD4^+^ count than their male counterparts, as shown in Table 1.

**Table 1.**
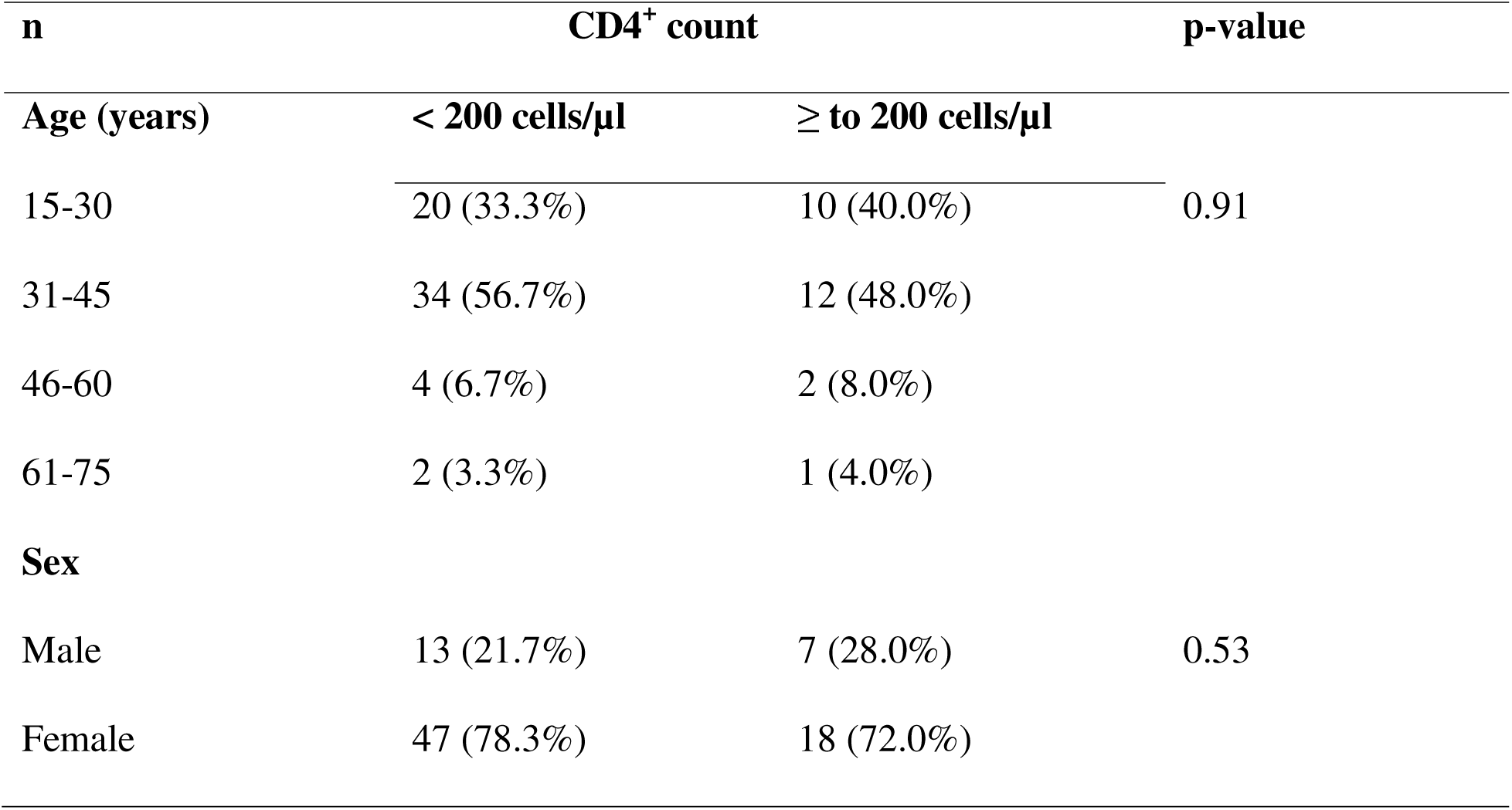
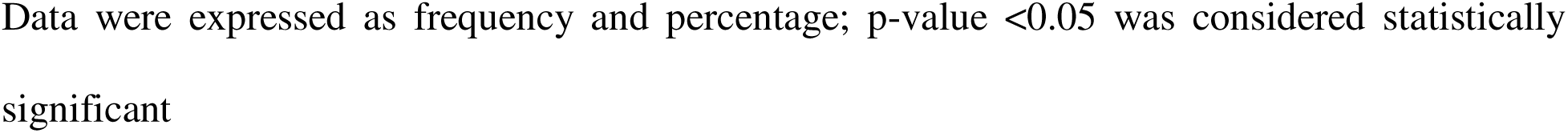
Demographic details of study participants with respect to CD4^+^ count levels.

| **n** | **CD4<sup>+</sup> count** |  | **p-value** |
| --- | --- | --- | --- |
| **Age (years)** | **< 200 cells/μL** | **≥ to 200 cells/μL** |  |
| 15-30 | 20 (33.3%) | 10 (40.0%) | 0.91 |
| 31-45 | 34 (56.7%) | 12 (48.0%) |  |
| 46-60 | 4 (6.7%) | 2 (8.0%) |  |
| 61-75 | 2 (3.3%) | 1 (4.0%) |  |
| **Sex** |  |  |  |
| Male | 13 (21.7%) | 7 (28.0%) | 0.53 |
| Female | 47 (78.3%) | 18 (72.0%) |  |

Data were expressed as frequency and percentage; p-value <0.05 was considered statistically significant

### 3.2 Diagnostic efficacy of LF-LAM test for diagnosing TB

In this study, lipoarabinomannan (LAM) reactivity was 29 of 85 (34.1%) and non-reactivity in 56 of 85 (65.9%) HIV-positive patients. Lipoarabinomannan reactivity was higher in patients with a CD4^+^ count < 200 cells/µl, 22 (75.9%), as opposed to 7 (24.1%) for those with a CD4^+^ count ≥ 200 cells/µl. For patients with a CD4^+^ count < 200 cells/µl, 38 (67.9%) were LAM non-reactive, as compared to 18 (32.1%) for those with a CD4^+^ count ≥ 200 cells/µl, p = 0.76. The majority of the patients with a CD4^+^ count < 200 cells/µl, were LAM-reactive, as compared to those with a CD4^+^ count ≥ to 200 cells/µl (75.9% vs 24.1%). We also observed that most of the patients that were LAM non-reactive also had a lower CD4^+^ count (67.9% vs 32.1%). There was no statistically significant difference in the proportion of reactive LAM results according to CD4^+^ count, as shown in Table 2.

**Table 2.** TB LAM reactivity in relation to CD4^+^count levels of study participants.

| CD4<sup>+</sup> COUNT | LAM reactive | LAM non-reactive | p-value |
| --- | --- | --- | --- |
| < 200 cells/μl | 22 (75.9%) | 38 (67.9%) | 0.76 |
| ≥ to 200 cells/μl | 7 (24.1%) | 18 (32.1%) |  |
| **Total** | **29** | **56** |  |

Data were expressed as frequency and percentage; p-value <0.05 was considered statistically significant; LAM=Lipoarabinomannan

### 3.3 Recency assay outcomes to LAM testing

Based on recency testing, 76 (89.4%) HIV-positive patients had long term infection, and 9 (10.6%) patients were diagnosed with a recent infection. In this study, TB LAM was reactive in 27 of 85 (31.7%) HIV-positive patients with long-term HIV infection and non-reactive in 49 (57.6%). Most of the patients with TB LAM non-reactive results had HIV for a long period of time and may have been on anti-retroviral therapy (ART) over a longer duration, while treatment may also be interrupted. TB LAM was reactive in 2 (2.4%) HIV-positive patients with recent HIV infection and non-reactive in 7 (8.2%), p = 0.42. It also applies that individuals with recent infection were correlated with a LAM non-reactive result. There was no statistically significant difference in the proportion of reactive LAM tests based on recency status, as shown in Table 3.

**Table 3.** TB LAM reactivity in relation to HIV recency status.

| **Recency** | **LAM reactive** | **LAM non-reactive** | **p-value** |
| --- | --- | --- | --- |
| Long term infection | 27 (31.7%) | 49 (57.6%) | 0.42 |
| Recent infection | 2 (2.4%) | 7 (8.2%) |  |
| **Total** | **29** | **56** | **85** |

Data were expressed as frequency and percentage; $p$ -value $<0.05$ is considered statistically significant; LAM=Lipoarabinomannan

## 4 DISCUSSION

One of the main causes of sickness and death worldwide still remains tuberculosis [16]. Because of this silent killer impact on global health, modelling studies suggest that a rapid, moderately sensitive test using an easily obtainable non-sputum sample is useful in changing the status quo [22,23]. Among all the biological samples used in disease diagnosis, urine as a diagnostic tool has shown much importance in the diagnosis of various diseases. Urine contains different biochemical, immune, and haematological parameters, including a variety of mycobacterial antigens, the most promising of which has been shown to be the cell wall lipopolysaccharide known as lipoarabinomannan [24,25].

To the best of our knowledge, this is the first study to assess the usage of urine LF-LAM in PLWH in the study area. This study examined preliminary findings on the use of the LAM test for the screening of tuberculosis among a population of 85 HIV-positive patients in relation to their CD4^+^ count. While earlier studies have investigated the use of diagnostic approaches such as genexpert, culture, etc. for the diagnosis of TB in HIV patients. However, less invasive and technically simple approaches like LAM Ag assay have not received full recommendation as a flexible option in poorly developed endemic areas. Moreover, most of the studies were conducted in tertiary health settings with a large sample size. However, this study centres on grassroots facilities, where advanced techniques might not be used in the early diagnosis of tuberculosis; due to cost, adequate power supply, expert demands, and many more.

Out of all the 85 study participants included, most of the HIV-positive patients had a CD4^+^ count < 200 cells/µl (70.6%). Those within the age group 31-45 had the highest percentage (56.7%) as compared to other age groups with a CD4^+^ count less than 200 cells/µl. The burden of a CD4^+^ count lower than the normal range was observed more in females in comparison than in their male counterparts. This might be due to the influence of homeostatic/hormonal imbalance, which could also fuel immunosuppression in females. CD4^+^ cells are important immune cells that play a crucial role against viral infections, including HIV. As a marker of immune function, when the immune system deteriorates, it also affects its action as the virus attacks it. There was also no statistically significant difference between the CD4^+^ count and independent variables like age and sex.

In this study, the reactivity of determine TB Ag LAM was 34.1% and non-reactive, 65.9%, respectively. Antigenemia in HIV/TB co-infected patients which is consequently filtered into urine, could be related to a higher bacterial burden [26], regardless of their ART status. Most of the non-reactive results were observed in patients with a lower CD4^+^ count. Reports showed that the sensitivity of LF-LAM is not accurate in individuals with a functioning immune system, but it is more effective in detecting MTB infections in individuals with HIV, especially those with a CD4+ count of less than 200 cells/µl [27]. In research by Huerga and colleagues, LAM positivity was 11.9% among symptomatic patients [28]. Another study revealed that urinary LAM had a percentage of 95.7% in TB patients but with the use of ELISA instead [29].

Based on the WHO recommendation, the LF-LAM test is not strongly advised for diagnosing tuberculosis in people living with HIV who have a CD4+ count higher than 200 cells/µL [30]. However, we included HIV-positive patients with CD4^+^ greater than 200 cells/µL in an attempt to simulate the usefulness of LAM testing in diagnosing tuberculosis in HIV-negative patients and other disease groups. Despite this, LF-LAM test diagnostic accuracy in HIV-negative patients from previous studies was not satisfactory, with reported estimated sensitivities ranging from 4% to 31% [31–34]. To improve health coverage, there is a need for more validated research on HIV-negative patients.

A meta-analysis of 15 studies conducted in 1761 TB patients with over 6800 participants documented the pooled sensitivity and specificity to be 42% and 91%, respectively [30]. 65% and 89.36% sensitivity and specificity, were reported in another study [35]. Other studies described a connection with being HIV positive, having a low CD4 count and an accompanied rise in LAM sensitivity [30, 36]. However, in this investigation, we did not find any correlation between low CD4 count and LAM reactivity in terms of sensitivity and specificity. One must exercise caution when interpreting this, as the study lacked enough statistical power to evaluate such a correlation [37], because there might be many false positive and false negative results, which might prevent us from ruling out TB completely. However, the small sample size might have contributed to this contrast in the LAM assay used as a standalone diagnostic test. Another study indicated that the Fujifilm SILVAMP TB-LAM test demonstrated significantly higher sensitivity in tuberculosis detection in people living with HIV compared to the LF-LAM assay. [38]. Further efforts are required to substantiate LF-LAM testing as an ideal bedside point-of-care diagnostic.

The outcome of this study highlighted that due to the limited financial resources available in affording important diagnostic testing for HIV-associated disease such as molecular testing, blood tests, chest X-ray; the use of LAM antigen testing for TB screening is indicative of its diagnostic resourcefulness in primary healthcare settings. It could also orchestrate a positive societal impact. Even though geneXpert is free, transporting sputum samples to reference laboratories could delay TB diagnosis [39], as it is not readily accessible to all healthcare facilities, especially the grassroots. Previous studies have also shown that sputum-based TB testing alone is not the best diagnostic and therapeutic approach in the HIV population, due to the inability to produce an adequate sputum sample for diagnosis [28,40]. Due to the anxiety, pain, and trauma associated with blood sample collection for diagnosis, the Alere LAM intervention could change the narrative in patient compliance for TB testing and management.

There are some contraindications to using LAM. First, there are cross-reactions between non-tuberculous mycobacteria (NTM) and LAM. This is because LAM does not distinguish between different species of Mycobacterium, such as *M. avium*, *M. tuberculosis*, M. *leprae*, *M. bovis*, and *M. ulcerans* [41,42]. It is also possible that people with advanced HIV disease may be more likely to have a non-tuberculous mycobacterial infection instead of tuberculosis. A significant number of HIV persons (19 out of 21) with disseminated NTM infection had a very high outcome of false-positive results in previous research [41]. Furthermore, the LAM urine assay employs polyclonal anti-LAM antibodies to capture the urinary LAM. This implies that it possesses numerous common epitopes between tuberculosis (TB) and non-tuberculous mycobacteria (NTM) [33]. Subsequent studies on the impact of NTM infection on LAM positivity are required to curb the global health burden of the disease.

Interestingly, the use of LAM testing in our study setting was cost-effective, required minimal logistics, had a short turnaround time, and was simple to perform with less technical expertise and non-invasive sample usage. Although WHO proposes LF-LAM as an adjunctive diagnostic test for HIV/TB coinfection in patients exhibiting TB symptoms/signs, advanced illness, and CD4+ T-cell counts < 200 cells/µl [43], most research works do not capture the demographic distribution in under-resourced facilities, where development is at its poorest. This study explains the need for more LF-LAM consumables in health facilities.

The strengths of this study were the detection of LAM Ag in urine samples from the study participants. Subsequently, we used fresh, unprocessed urine samples, as it is noted that fresh urine samples contain more detectable LAM. This might be more crucial in patients with a CD4 count higher than 200 cells/µL with the likelihood of low LAM concentrations [44,45].

### 4.1 Limitations of the study

However, we acknowledge the following limitations: (1) The sample size is small, potentially insufficient to draw strong conclusions about the diagnostic tool’s effectiveness in early TB detections in highly burdened areas; (2) limited to only HIV-positive patients; (3) TB prevalence was not assessed in the overall population; (4) absence of other TB diagnostic testing such as geneXpert to detect rifampicin resistance, smear/culture; and (5) we did not correlate the viral load (VL) level of the HIV-positive patients.

Therefore, we recommend further research incorporating non-HIV populations with the use of other TB diagnostic algorithm to validate the sensitivity and specificity of LAM. Assessing LAM Ag in children rather than adults alone will help initiate a wide coverage of its importance and more informed clinical decisions. In addition, a larger sample size is needed to strengthen the findings and make effective contributions to the adoption of LF-LAM in most healthcare facilities across TB endemic areas in Nigeria and Africa.

## 5 CONCLUSION

Putting an end to the health burden of infectious diseases like TB has become a global effort. The findings from this study suggest that urinary LAM assay could serve as a solution-driven option to addressing the challenge of the late detection of TB in HIV in resource-deprived settings. Urine LAM Ag assay implementation would not only lower diagnostic costs and promote early initiation of TB treatment, but also reduce the frequent requirement for a steady power supply, which is vital for both smear and culture procedures, including advanced PCR techniques in rural settings. This promising diagnostic tool is on the path to revolutionising TB treatment and management outcomes.

## Data Availability

All data produced in the present study are available upon reasonable request to the authors

## DECLARATIONS

### Ethics approval and consent to participate

Informed consent and approval was obtained from participants and GHUA Medical Ethics Committee of the healthcare facility, Ministry of Health, Nigeria.

### Consent for publication

Not applicable.

### Availability of data and materials

Data will be made available to interested parties on reasonable request

### Authors’ contributions

AIA, GB, SEG, BDO and RY conceived and designed the study. AIA and GB wrote the main part of the Introduction, Methodology and Discussion sections. SEG arranged the bibliography and created figures. RY, SEG and BDO contributed to the introduction and discussion sections. AIA and BDO collected the data. NE reviewed the draft of the manuscript and study implementation. AIA analysed the data and GB reviewed the analysis. AIA wrote the conclusion part. All authors read and approved the final manuscript.

## Acknowledgements

We are grateful to Mohammed F Alawami, ReachSci Society, for reviewing the draft of the manuscript, supporting the study and for providing constructive guidance throughout the study timeline.

## List of abbreviations

ART: Anti-retroviral therapy
CD4: Cluster of Differentiation
ELISA: Enzyme-Linked Immunosorbent Assay
DR: Drug resistant
FEND TB: Feasibility of Novel Diagnostics of TB in Endemic Countries
HIV: Human Immunodeficiency Virus
LAM: Lipoarabinomannan
LF: Lateral flow
MTB: *Mycobacterium tuberculosis*
NIAID: National Institute of Allergy and Infectious Diseases
NTM: Non-tuberculous Mycobacteria
PCR: Polymerase Chain Reaction
PLWH: People Living with HIV
SPSS: Statistical Package for Social Science
TB: Tuberculosis
VL: Viral Load
WHO: World Health Organization

